# Arabic validation and cross-cultural adaptation of the 5C scale for assessment of COVID-19 vaccines psychological antecedents

**DOI:** 10.1101/2021.02.03.21251059

**Authors:** Samar Abd ElHafeez, Ramy Shaaban, Iffat Elbarazi, Rony ElMakhzangy, Maged Ossama Aly, Amr Alnagar, Mohamed Yacoub, Haider M. El Saeh, Nashwa Eltaweel, Sulafa T Alqutub, Ramy Mohamed Ghazy

## Abstract

**Background:** In the Arab countries, there has not been yet a specific validated questionnaire that can assess the psychological antecedents of COVID-19 vaccine among the general population. This study, therefore, aimed to translate, culturally adapt, and validate the 5C scale into the Arabic language.

**Methods:** The 5C scale was translated into Arabic by two independent bilingual co-authors, and then subsequently translated back into English. After reconciling translation disparities, the final Arabic questionnaire was disseminated into four randomly selected Arabic countries (Egypt, Libya, United Arab Emirates (UAE), and Saudi Arabia). Data from 350 Arabic speaking adults (aged ≥18 years) were included in the final analysis. Convergent, discriminant, exploratory and confirmatory factor analyses were carried out. Internal consistency was assessed by Cronbach alpha.

**Results:** Age of participants ranged between 18 to 73 years; 57.14% were females, 37.43% from Egypt, 36.86%, from UAE, and 30% were healthcare workers. The 5 sub-scales of the questionnaire met the criterion of internal consistency (Cronbach alpha ≥0.7). Convergent validity was identified by the significant inter-item and item-total correlation (*P*<0.001). Discriminant validity was reported as inter-factor correlation matrix (<0.7). Exploratory factor analysis indicated that the 15 items of the questionnaire could be summarized into five factors. Confirmatory factor analysis confirmed that the hypothesized five-factor model of the 15-item questionnaire was satisfied with adequate psychometric properties and fit with observed data (RMSEA=0.060,GFI=0.924, CFI=0.957, TLI=0.937, SRMR=0.076 & NFI=906).

**Conclusion:** the Arabic version of the 5C scale is a valid and reliable tool to assess the psychological antecedents of COVID-19 vaccine among Arab population.

## Introduction

The world is currently in a public health crisis facing a fierce virus, the Coronavirus (COVID-19), which puts the world in a pandemic. COVID-19 spreads rapidly around the globe, affecting more than one hundred million with a toll of death exceeding two million within one year [1].

All the countries around the world are fighting the spread of COVID-19. Procedures that countries have taken include enforcing quarantines, lockdowns, social distancing, wearing facemasks, and travel restrictions. These procedures have impacted people both physically and psychosocially and have massively left negative impacts on the global economy. “The multi-faceted catastrophic consequences associated with the COVID-19 outbreak have intensified international efforts in developing an effective prevention method to keep outbreaks under control” [2].

A combined effort is being simultaneously exerted by the World Health Organization (WHO), governments, academic community, and pharmaceutical industries to develop and deploy safe and effective vaccines. To this date, more than 100 vaccines are in a pre-clinical development phase, with at least 50 vaccines selected to reach the clinical development stage [3]. On 31 December 2020, the WHO listed mRNA vaccine (Pfizer/BioNTech) for emergency use [4].

The production of an effective vaccine against COVID-19 virus faces several challenges such as selecting a proper formulation, reviewing and approving a large number of potential vaccine candidates, massively producing the vaccine, and surveilling it in the post-marketing stage, in addition to the cost issues and logistics of distribution [5-7]. Nevertheless, a major obstacle towards achieving appropriate vaccination and reaching an eventual herd immunity can be vaccine hesitancy among the general public. Newly emerging vaccines are usually questioned by community members and the views on receiving them can vary dramatically between individuals [8].

Vaccine Hesitancy (VH) refers to postponing or refusing to accept vaccination despite its availability [9]. It is a complex situation that varies across time, place and type of the vaccine. Vaccine hesitancy has become a major obstacle in the face of the preventive strategies and procedures that aim to fight the spread of infectious diseases and is predicted to slow the fight against the prospective COVID-19 vaccines [10]. In addition, solely depending on the vaccination can lead into an un-favorite outcome if other protection strategies are neglected [11].

There is a significant difference in vaccine acceptance. In a global survey conducted by Lazarus V *et al* [12]; 71.5% responded that they would accept and take the vaccine in case it were proven safe and effective, and 48.1% said that they would get vaccinated if their employer suggested it. Moreover, a worldwide systematic review on COVID-19 vaccine acceptance showed that the highest acceptance rates of COVID-19 vaccination were found in Ecuador (97.0%), Malaysia (94.3%), Indonesia (93.3%) and China (91.3%), respectively. On the opposite side, the lowest acceptance rates of COVID-19 vaccination were found in Kuwait (23.6%), Jordan (28.4%), Italy (53.7), Russia (54.9%), Poland (56.3%), US (56.9%), and France (58.9%), respectively[13].

Several questionnaires were developed to assess the vaccine acceptance and hesitancy, as Vaccine Confidence Scale [14], Parent Attitudes about Childhood Vaccines Survey [15], Vaccine Hesitancy Scale (VHS) [16], Global Vaccine Confidence Index [17], and the 5C scale [18].

The 5C scale extends “the scope of available measures and covers the broader theoretical conceptualization of vaccine hesitancy and acceptance” [2]. It measures the five psychological antecedents or determinants within the individual that is related to whether or not he/she vaccinates. In addition, “it provides insights into the individual mental representations, attitudinal and behavioural tendencies that are a result of the environment and context the respondent lives in” [19]. These five antecedents are confidence, complacency, constraints, calculation, and collective responsibility [19].

In the Arab region, two studies were conducted to assess the COVID-19 vaccine hesitancy [20, 21]. Both studies did not report using an Arabic validated questionnaire. This study, hence, aimed to translate, culturally adapt, and validate the long form of the 5C questionnaire into the Arabic language.

## Methods

### The study tool

The 5C scale is composed of five sub-scales assessing the different psychological antecedents of vaccination [19]. It includes:

a. Confidence: is defined as “trust in (i) the effectiveness and safety of vaccines, (ii) the system that delivers them, including the reliability and competence of the health services and health professionals, and (iii) the motivations of policy-makers who decide on the need of vaccines” [9].
  - I am completely confident that vaccines are safe’’Q1’’.
  - Vaccinations are effective “Q2”.
  - Regarding vaccines, I am confident that public authorities decide in the best interest of the community “Q3”.
b. Complacency: “exists where perceived risks of vaccine-preventable diseases are low and vaccination is not deemed a necessary preventive action” [9].
  - Vaccination is unnecessary because vaccine-preventable diseases are not common anymore”Q4”.
  - My immune system is so strong, it also protects me against diseases “Q5”.
  - Vaccine-preventable diseases are not so severe that I should get vaccinated”Q6”.
c. Constraints: are an issue when “physical availability, affordability and willingness-to-pay, geographical accessibility, ability to understand (language and health literacy) and appeal of immunization service affect uptake” [9].
  - Everyday stress prevents me from getting vaccinated “Q7”.
  - For me, it is inconvenient to receive vaccinations”Q8”.
  - Visiting the doctor’s makes me feel uncomfortable; this keeps me from getting vaccinated”Q9”.
d. Calculation: refers to individuals’ engagement in extensive information searching. It is related to perceived vaccination and disease risks [22].
  - When I think about getting vaccinated, I weigh benefits and risks to make the best decision possible “Q10”.
  - For each and every vaccination, I closely consider whether it is useful for me “Q11”.
  - It is important for me to fully understand the topic of vaccination, before I get vaccinated “Q12”.
e. Collective responsibility: it is the willingness to protect others by one’s own vaccination by means of herd immunity [23].
  - When everyone is vaccinated, I don’t have to get vaccinated, too “Q13”.
  - I get vaccinated because I can also protect people with a weaker immune system “Q14”.
  - Vaccination is a collective action to prevent the spread of diseases “Q15”.

All items were rated on 7-point Likert scale (1 = strongly disagree, 2 = moderately disagree, 3 = slightly disagree, 4 = neutral, 5 = slightly agree, 6 = moderately agree, and 7 = strongly agree).

### Score interpretation

Because a general term that contains all antecedents does not exist, it is not theoretically acceptable to calculate a total score for all antecedents. Using the 5C scale does not lead to a total score providing a sample’s absolute state of hesitancy. It, rather, allows for a valid assessment of determinants and predicts vaccination. It, therefore, allows intervention design informed by monitoring and evidence practices [19, 24].

### Translation and adaptation

We forward-translated the 5C scale into Arabic by two independent bilingual co-authors (AA & NE). Both co-authors rated the difficulty of translating each item and the associated response choices. One bilingual researcher (RS) and another Arabic translator compared the two translations and reconciled the discrepancies. Then, the questionnaire was back translated into English by two additional co-authors (MY & RE). The back translators compared their translations with the previous English version. Minor discrepancies were identified and resolved by discussions between the researchers.

### Content validity and expert evaluation

The next step in the validation process was to assess the content validity with an expert panel of 10 investigators (methodologist, healthcare professionals, public health professional, and language professionals). The expert panel examined whether the agreed-on translation covers the concepts as defined.

### Cognitive interviews

We next performed cognitive testing of the Pre-final version. Trained members of the research team conduced cognitive interviews among 20 participants of the intended respondents (5 from each included country) to evaluate participant understanding, readability, language, wording, and cultural appropriateness of items as well as the clarity of the instructions for providing responses for each section.

During this step, we encountered some difficulties with explaining some points. The first comment was related to the seven points Likert scale, particularly the difference between strongly agree/disagree and moderately agree/disagree. In the Arabic language, there is no sharp demarcation between the perceived meaning of strongly and moderately. Another item, which was not well understood by the participants, is the “Everyday stress prevents me from getting vaccinated”, there was a confusion regarding the real perspective of the daily stress will hinder them from taking the vaccine. Some participants felt that there was a repetition of the questions “Vaccination is unnecessary because vaccine-preventable diseases are not common anymore” and “Vaccine-preventable diseases are not so severe that I should get vaccinated”. We reformulated the Arabic questions to deliver the construct beyond each item of the original copy of the questionnaire. Then, the final Arabic version was approved by the researchers and ready for field testing.

### Data set for testing the validity of the Arabic version of 5C scale

Based on the sample size recommendations of having 10 participants respond to each item for validating a questionnaire (ratio 10:1), we needed 150 participants [25]. Moreover, a priori sample size calculation for Structural Equation Modelling (SEM) technique to perform confirmatory factor analysis (CFA) showed that a minimum sample of 200 is required to run CFA [26]. For that, the minimum required sample size for our analysis was 350 participants. Adult (18 years and above) Arabic speaking population is included in the study.

The final Arabic copy of 5C scale was uploaded on Qualtrics and disseminated online via different social media platforms (Facebook, WhatsApp, emails, and Twitter) to 673 participants. The sample was recruited from four randomly selected Arabic countries (Egypt, Saudi Arabia. Libya, and United Arab of Emirates (UAE)). A total of 511 responded to the questionnaire, 89 participants chose not to complete the questionnaire. The response rate was 62.70% (422/673). Of the 422 who completed the questionnaire, we excluded 72 responses from the final analysis due to incomplete or inconsistent data. The final sample size included in our analysis was 350 participants.

### Data management and psychometric analysis

Quantitative data are summarized as mean ± standard deviation (SD) while qualitative data are presented with percent and frequency. Total scores of each sub-scale were calculated by addition of the scores of all items within each sub-scale. Pearson’s correlation analysis was used to calculate inter-item and item-to-total.

### Reliability and Item Analysis

Cronbach’s alphas were calculated for the sub-scales of the questionnaire to assess its internal consistency. As a rule of thumb, a Cronbach’s alpha of 0.70 to 0.80 is considered respectable for a scale for research use and an alpha more than 0.80 is considered very good [27].

### Construct Validity

it represents the “extent to which an instrument assesses a construct of concern, and is associated with evidence that measures other constructs in that domain and measures specific real-world criteria” [28]. It is determined using content, criterion-related validity, and structural or factorial validity [28].

#### Criterion-related validity

both convergent and discriminant (divergent) validity were used as indicators of criterion-related validity. Convergent validity was assessed by analyzing inter-item and item-to-total correlation. Discriminant validity was assessed by calculating factor correlation matrix of the five subscales[29].

#### Factorial analysis validity

We analyzed data collected from 350 participants. Factor analysis was performed in two steps: exploratory and confirmatory factor analysis (EFA and CFA). We randomly divided the participants into two groups; 150 participants for EFA and 200 participants for CFA.

### Exploratory Factor analysis

The EFA aimed at identifying the major factor structures for the set of 15 items and to determine the number of latent factors, without making assumptions about the factor relationships [30]. Kaiser-Meyer-Olkin (KMO) sampling adequacy measure and Bartlett’s sphericity test were performed before EFA. The KMO statistics range from 0 to 1, with values closer to 1 denoting greater adequacy of the factor analysis (KMO ≥ 0.6 low adequacy, KMO ≥ 0.7 medium adequacy, KMO ≥ 0.8 high adequacy, KMO ≥ 0.9 very high adequacy) and P value of Bartlett’s test is < 0.05, then factorial analysis can be used [31]. The number of factors extracted is based on Eigenvalues (>1), scree plot, parallel analysis, and interpretability of the factors [32].

To determine the type of rotation, we first ran EFA using the principal component analysis with the an oblique direct Oblimin rotation to calculate the inter-factor correlation. Discriminant validity was assessed if inter-factors correlation based on the factor correlation matrix is less than 0.7 [33].

The final EFA was done using the principal component analysis with the orthogonal Varimax rotation. A factor loading cut-off value of 0.50 was chosen to decide which items were highly associated with a given factor[32]. In interpreting the output, we defined that each factor should have at least 3 items with high factor loadings of 0.5 and higher on the primary factor and minimal cross-loadings on any of the other factors (a < 0.35) to reduce the overlap between the sub-scales [32, 34].

### Confirmatory factor analysis

The CFA that was performed based on the selected 200 participants aimed to measure how well the factor structure, identified in the EFA, fits the observed data. Specifically, we assessed the convergent and discriminant validity of the constructs and model fit measures using the SEM technique [35]. We used the root mean square error of approximation (RMSEA <0.08), comparative fit index (CFI >0.9), Tucker Lewis index (TLI>0.9), standardized root means square residual (SRMR ≤0.08), normal fit index (NFI>0.9), goodness of fit (GFI>0.9) as model fit indicators, and χ^2^/df <3 [36]. Convergent validity was determined if the average variance extracted (AVE) values of the different factors were above 0.5. Discriminant validity was confirmed if the square root of AVE is higher than the inter-correlation between the factors[37]. Moreover, we assessed the construct reliability of each latent factor and reliability ≥0.7 indicates good reliability [37]. We used statistical package of social science SPSS (version 25, Chicago, USA) and SPSS AMOS 26 to run all the analyses.

### Ethical considerations

The study was approved by the Ethics Committee of the Faculty of Medicine-Alexandria University, Egypt (IRB No:00012098) following the International Ethical Guidelines for Epidemiological studies [38].

## RESULT

### Characteristics of the study participants

Table 1 shows the baseline characteristics of the study population. Age ranged between 18 to 73 years; mean age of 34 ± 12 years. More than half were females (57.14%), 37.43% were from Egypt, and 36.86 were from UAE. One-third were healthcare workers and one-half (51.14%) were university graduates. Only 16.29% reported a previous history of COVID-19 infection, 38.57% gave a family history of death due to the infection, and 79.42 % reported knowing about the several types of vaccines.

**Table 1:** Baseline characteristics of study population.

| **Baseline characteristics** | **Frequency (%)**<br>**(N=350)** |
| --- | --- |
| **Age** |  |
| 18-30 | 104(29.71) |
| 31-45 | 149(42.57) |
| 46-60 | 71(20.57) |
| >60 | 25(7.14) |
| **Mean± SD age in years** | 34 ± 12 |
| **Sex** |  |
| Male | 150(42.86) |
| Female | 200(57.14) |
| **Country** |  |
| Egypt | 131(37.43) |
| Libya | 34(9.71) |
| United Arab of Emirates | 129(36.86) |
| Saudi Arabia | 56(16.00) |
| **Education** |  |
| Secondary | 48(13.71) |
| Vocational education | 18(5.14) |
| University graduate | 179(51.14) |
| Post-graduate | 99(28.29) |
| Others | 6 (1.71) |
| **Chronic diseases** |  |
| Yes | 75(21.4) |
| No | 275(78.57) |
| **Health care workers** |  |
| Yes | 105(30.00) |
| No | 245(70.00) |
| **Did you get COVID-19 infection** |  |
| Yes | 57(16.29) |
| No | 225 (64.29) |
| I do not know | 68(19.42) |
| **If there any of your relative died due to COVID-19 infection** |  |
| Yes | 135(38.57) |
| No | 215(61.43) |
| **Do you know that there is many types of COVID-19 vaccine** |  |
| Yes | 278 (79.42) |
| No | 72(20.58) |

### Questionnaire validation

We ran univariate item analysis using collected data from 150 participants. All items means ranged from a minimum of 2.17 to a maximum of 6.14, and SD ranged from 1.25 to 1.94. Table 2 shows the descriptive statistics of the different items of the questionnaire (Table 2).

**Table 2:** Descriptive statistics, reliability, and convergent validity of the Arabic version of the 5C scale.

| Variable | Mean ± SD | Item-total correlation |
| --- | --- | --- |
| **Confidence** |  |  |
| Q1 | 4.65±1.73 | 0.91($P < 0.001$) |
| Q2 | 4.93±1.57 | 0.87($P < 0.001$) |
| Q3 | 5.15±1.92 | 0.82($P < 0.001$) |
| **Cronbach's alpha** | **0.829** |  |
| **Complacency** |  |  |
| Q4 | 2.17±1.79 | 0.81($P < 0.001$) |
| Q5 | 3.76±1.86 | 0.79($P < 0.001$) |
| Q6 | 3.07±1.94 | 0.79($P < 0.001$) |
| **Cronbach's alpha** | 0.712 |  |
| **Constraints** |  |  |
| Q7 | 2.81±1.77 | 0.70($P < 0.001$) |
| Q8 | 3.12±1.79 | 0.82($P < 0.001$) |
| Q9 | 2.75±1.85 | 0.79($P < 0.001$) |
| **Cronbach's alpha** | 0.701 |  |
| **Calculation** |  |  |
| **Q10** | 5.51±1.67 | 0.84($P < 0.001$) |
| **Q11** | 5.76±1.39 | 0.86($P < 0.001$) |
| **Q12** | 6.14±1.36 | 0.80($P < 0.001$) |
| **Cronbach's alpha** | 0.773 |  |
| **Collective responsibility** |  |  |
| **Q13** | 5.85±1.25 | 0.80($P < 0.001$) |
| **Q14** | 5.60±1.73 | 0.91($P < 0.001$) |
| **Q15** | 6.03±1.37 | 0.89($P < 0.001$) |
| **Cronbach's alpha** | 0.829 |  |

### Reliability analysis

all sub-scales had a satisfactory internal consistency. Both “Confidence” and “Collective responsibility” sub-scales have Cronbach’s alpha of 0.829.” “Constraints” sub-scale had the lowest Cronbach’s alpha (0.701). (Table 2)

### Convergent validity

inter-item correlation for each sub-scale was highly significant (P<0.001) (Table 1 supplementary). In addition, item-total correlation was significant. (Table 2)

### Exploratory factorial analysis

Before conducting the EFA, we assessed the sampling adequacy and sphericity assumptions. KMO measure of sampling adequacy was 0.80, which is above the recommended value of 0.60, and Bartlett’s test of sphericity was found to be highly significant (*P* < 0.001). Moreover, all the communalities demonstrated to be 0.5 or more.

Using these previously mentioned indicators, we conducted an EFA; at first, we ran the analysis in the form of principal component analysis with an oblique direct Oblimin rotation to assess the factor correlation matrix and check the discriminant validity. There were both negative and positive correlations among the five factors. The largest negative correlation was between Complacency and Constraints (−0.276), while the smallest negative correlation was between Complacency and Calculation (−0.074). The largest positive correlation was between Confidence and Constraints (0.300), while the lowest positive correlation was between Calculation and Collective responsibility (0.033). There were no correlation coefficients larger than 0.7; hence, the factors derived from EFA revealed adequate discriminant validity (See details in Table 3).

**Table 3.** Factor correlation matrix of the Arabic version of the 5C scale.

| **Factor** | Confiden<br>ce | Complac<br>ency | Constrai<br>nts | Calculati<br>on | Collective<br>responsibility |
| --- | --- | --- | --- | --- | --- |
| Confidence | 1.000 |  |  |  |  |
| Complacency | -0.208 | 1.000 |  |  |  |
| Constraints | 0.300 | -0.276 | 1.000 |  |  |
| Calculation | -0.077 | -0.074 | 0.226 | 1.000 |  |
| Collective<br>responsibility | -0.174 | 0.241 | -0.248 | 0.033 | 1.000 |

The final analysis took the form of the principal component analysis with Varimax rotation. The initial Eigenvalues showed that all 15 items of the questionnaire explained 72.8% of the variance in 5 factors. Table 4 shows the factor loadings for all items of the questionnaire. For “**Confidence sub-scale**,**”** the items were loaded on one factor with loading ranges from 0.782 to 0.868. For **the “Complacency sub-scale**,**”** all items were loaded on one factor with factor loading ranges from 0.736 to 0.793. For “**Constraints sub-scale**,**”** items loaded on one factor, with loadings from 0.606 to 0.861. For **“Calculation sub-scale**,**”** the items loaded on one factor, with loadings between 0.726 to 0.863. Lastly, for “**Collective responsibility**,**”** all items loaded on one factor with factor ladings ranges between 0.478 to 0.808.

**Table 4:** Factor loadings of the Arabic version of 5C scale.

| Items | Factor |  |  |  |  | Communalities |
| --- | --- | --- | --- | --- | --- | --- |
|  | Confidence | Complacency | Constraints | Calculation | Collective responsibility |  |
| **Q1** | **0.875** | -0.158 | -0.106 | -0.125 | 0.182 | 0.851 |
| **Q2** | **0.833** | -0.245 | -0.016 | -0.029 | 0.193 | 0.792 |
| **Q3** | **0.758** | 0.216 | -0.091 | 0.027 | 0.252 | 0.693 |
| **Q4** | -0.270 | **0.772** | 0.255 | -0.155 | 0.016 | 0.758 |
| **Q5** | 0.074 | **0.774** | 0.074 | 0.115 | -0.214 | 0.669 |
| **Q6** | -0.060 | **0.745** | 0.042 | 0.007 | -0.143 | 0.581 |
| **Q7** | 0.124 | 0.026 | **0.854** | -0.117 | 0.002 | 0.76 |
| **Q8** | -0.323 | 0.258 | **0.657** | 0.068 | -0.245 | 0.667 |
| **Q9** | -0.394 | 0.215 | **0.571** | 0.049 | -0.261 | 0.598 |
| **Q10** | -0.087 | 0.000 | 0.074 | **0.834** | -0.010 | 0.708 |
| **Q11** | 0.073 | -0.013 | -0.020 | **0.868** | 0.148 | 0.78 |
| **Q12** | -0.112 | 0.030 | -0.160 | **0.734** | 0.319 | 0.68 |
| **Q13** | 0.320 | -0.316 | -0.265 | 0.079 | **0.594** | 0.632 |
| **Q14** | 0.301 | -0.125 | -0.091 | 0.141 | **0.817** | 0.802 |
| **Q15** | 0.220 | -0.156 | -0.088 | 0.319 | **0.825** | 0.862 |

### Confirmatory Factor Analysis

To determine whether EFA proposed five-factor model with the 15-item questionnaire can be used as a valid tool towards assessment of the psychological antecedents of COVID-19 vaccines among the Arab population, we conducted a CFA using a different sample of 200 participants.

We ran the CFA on the 15 items. We described the results of the CFA final model with the SEM shown in Figure 1. All the loadings were from 0.41 to 0.94. The construct reliability of the five factors in the CFA final model were above 0.7. For convergent validity, the average variance extracted (AVE) values of **confidence, complacency** and **calculations** factors were above 0.5. Although the AVE value of **constraints and collective responsibility** factors were less than 0.5, the factors specific items loadings were acceptable for convergent validity since there were no items with loading below 0.4. The correlation between the five latent variables was less than squared root of AVE, hence no problem with discriminant validity.

**Figure 1:**
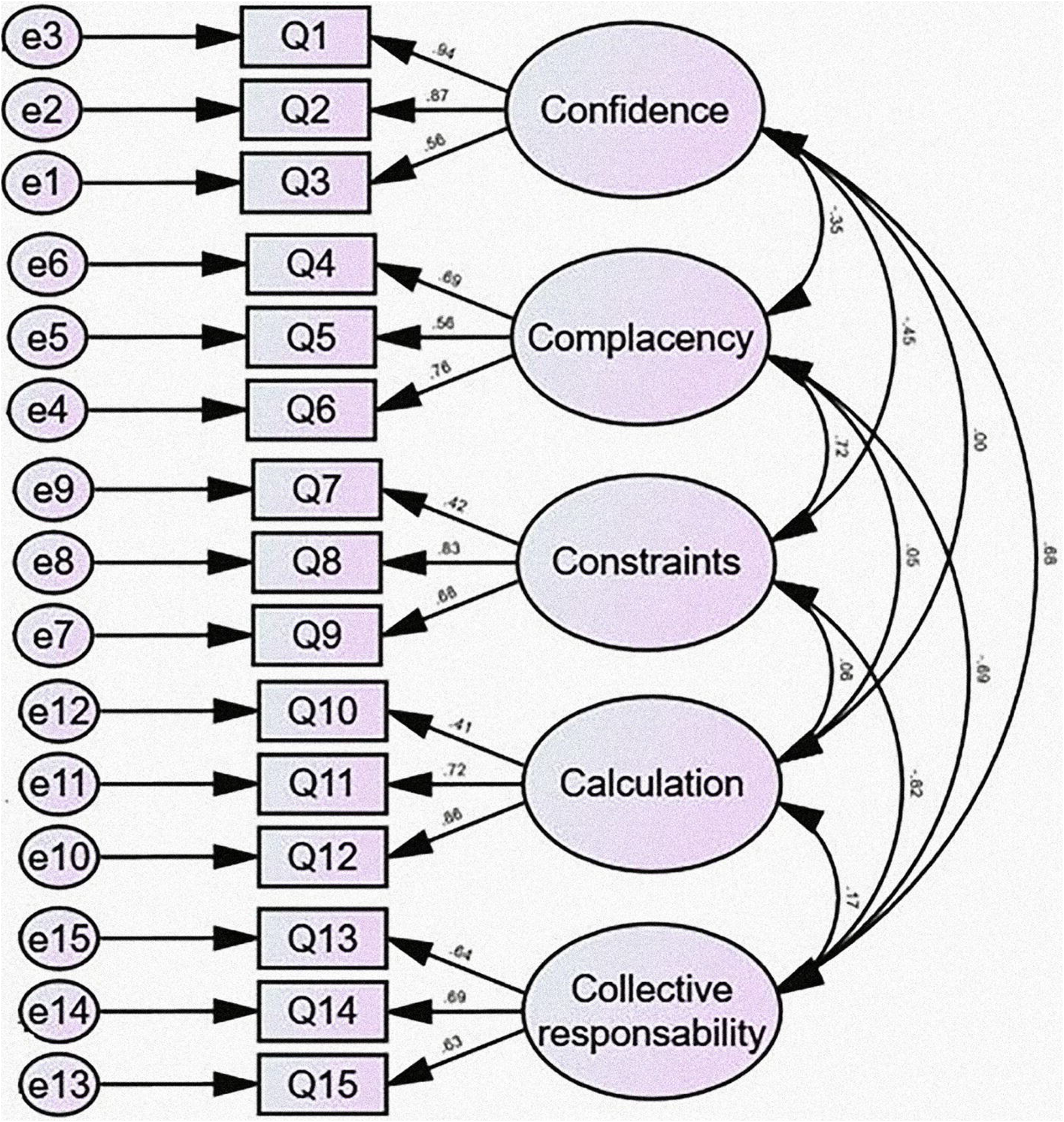
Confirmatory factor analysis of the 15 items of the Arabic version of the 5C scale.

An overview of goodness-of-fit measures for the final model is shown in table 5. The results demonstrate good model-data-fit, i.e., RMSEA <0.08, GFI, NFI, CFI, and TLI >0.9, and SRMR<0.08. Hence, the 15-item questionnaire has good psychometric properties and model fit to observed data.

**Table 5.**
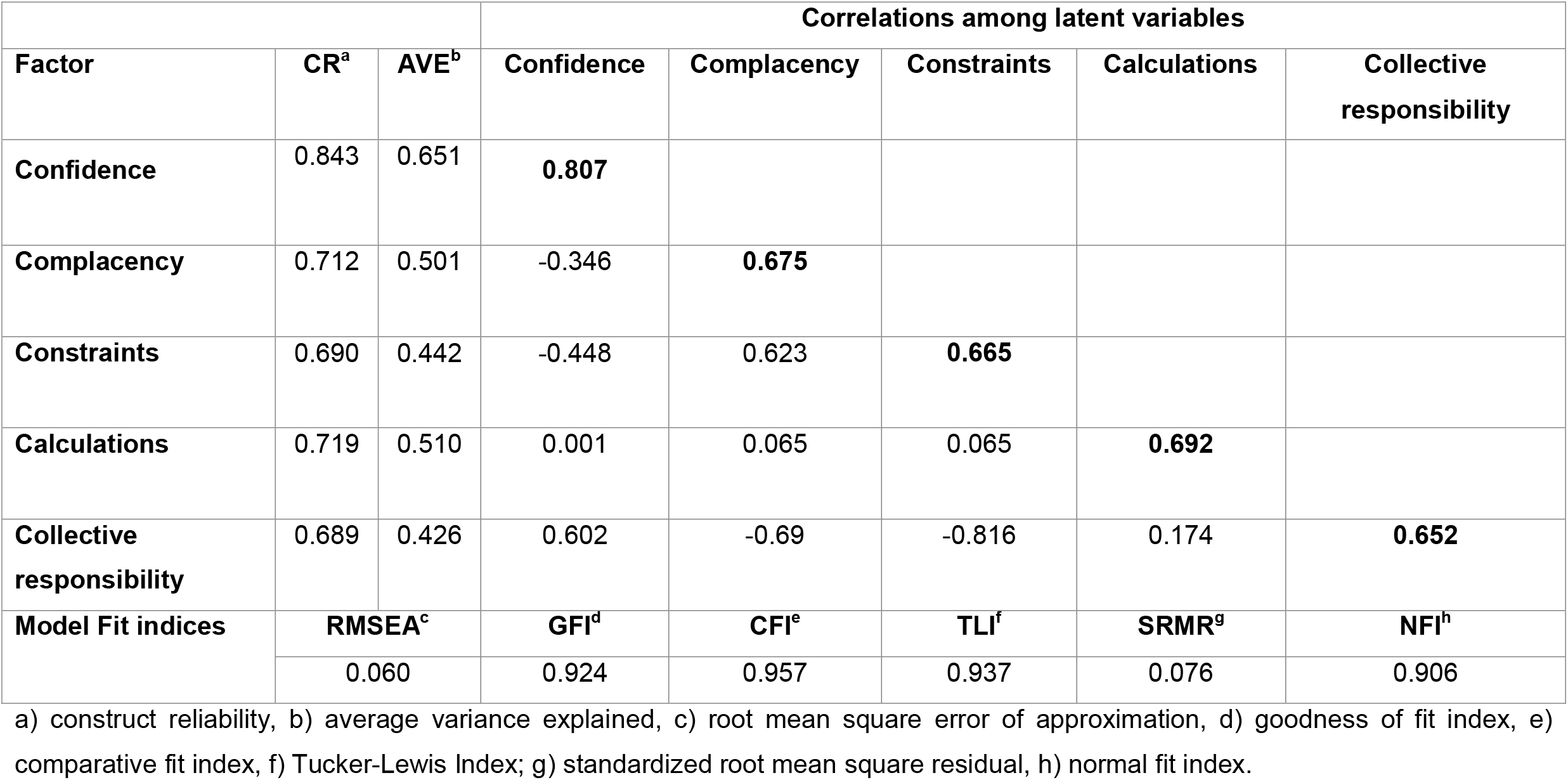
Results of the confirmatory factor analysis of 5C scale (15 items). Convergent validity, discriminant validity, and reliability assessment of CFA final model with five latent factors and model fit indices.

## Discussion

This study reports the results of the validation of the Arabic version of the 5C scale for assessment of the psychological antecedents for COVID-19 vaccines. The study sample was randomly recruited from four Arab countries Egypt, Libya, UAE, and Saudi Arabia.

Differences between regions, populations, and cultures require reliability and validity assessment of measurement instruments [39]. In the Arab countries, this is the first validation study of the 5C questionnaire to assess the COVID-19 vaccine antecedents. Arabic-speaking countries share similar culture and language. Although different dialects are used, formal Arabic is the official language regardless the geographical location. Therefore, chosen countries in this study are good representative of the Arab region.

Psychometric test results were close to the values of the corresponding items in the original German questionnaire [19]. Lower value of Cronbach’s alpha was obtained from constraints sub-scale (0.70) compared to the original questionnaire (0.85). On the other hand, the Arabic version of the questionnaire showed a higher Cronbach’s alpha (0.83) for the collective responsibility sub-scale compared to the original questionnaire (0.71). This may be explained by the different context in which we tested the 5C scale. While the original questionnaire was tested before the era of COVID-19 pandemic, our questionnaire was peculiarly validated for COVID-19 vaccines. The debates about the different vaccines efficacy and safety affect the population acceptance. In addition, the vaccines are not still available in all countries due to different polices regarding the eligibility and stock availability.

We did EFA to decide the number of factors needed to be extracted from the Arabic version of the 5C scale. The resulting factors loadings provided that the five-factor solution was the best for our data. This is consistent with what has been reported from the factor analysis of the original version of the questionnaire[19].

We tested the EFA proposed five-factor model by CFA using different samples. The results confirmed that the Arabic version of 5C scale constitutes five underlying constructs, i.e., confidence, complacency, constraints, calculations, and collective responsibility. These constructs can be measured with the 15-item questionnaire.

We assessed the convergent and discriminant validity of the Arabic version of the 5C scale during CFA based on the AVE values [40]. In our study, convergent validity was documented for all factors except for two latent factors (constraints and collective responsibility) that had AVE values below 0.5; however, all factors loading were greater than or equal to 0.4 and significant during CFA. Moreover, these items measured the critical aspect of the different constraints and collective responsibility in the respective domains. Hence, those factors were kept in the model. We also assessed the discriminant validity by comparing the square root of AVE with inter-factor correlation. Since square root of AVE was larger than the inter-factor correlation, the discriminant validity issues were not possible for the Arabic version of the 5 C scale. Among the five latent factors, the construct reliability was above the minimum acceptable level; which indicates a good internal consistency of the questionnaire.

The 5C scale can adequately and accurately assess the psychological foundations of vaccine acceptance and hesitancy. It is also indicative of the participants’ psychological statuses. An individual who lacks confidence is less likely to have a positive attitude and beliefs, less likely to trust the health system and medical treatments in general, and tends to accept conspiracy theories or take them for granted. An individual who is tied to constraints also tends to have a more general lack of self-control and self-efficacy. Highly constrained individuals perceive a lack of time. Therefore, vaccination should be made handy for them. A typical complacent person is not vulnerable; they feel healthy and tend to not care about their health future, which might lead them to act in a high-risk way. Disease risks are perceived low for this type of people. Conversely, people who calculate risks prefer to deliberate and are usually concerned about the risks associated with vaccination. Although deliberation and risk calculations are to consider, the respective skills (numeracy) are not especially high in these people, which can lead to skewed risk perceptions—i.e., high vaccination risks and low disease risks. “People who score high on collective responsibility generally care more for other people and are more empathic” [24]. Consequently, 5C scale is considered an optimal tool for different determinants measurement of vaccine acceptance.

Based on the findings of this study, the Arabic version of the 5C scale is a valid and reliable tool to assess the psychological antecedents of COVID-19 vaccine among Arabic speaking population.

## Data Availability

Data is available upon request by mailing the first author

## Author contribution

SA, RG conceptualized and designed the study. NE, AA, RE, RS, and MY participated in the 5C questionnaire translation. MO, RE, IE, HE, and STA performed the data collection, SA and RG analyzed and interpreted the data. SA, RG, and IE drafted and revised the manuscript critically. All authors read and approved the final manuscript.

## Data availability

Data is available upon request by mailing the corresponding author.

## References

1. Worldmeter. Corona virus 2020 [cited 2021 Jan, 29]. Available from: https://www.worldometers.info/coronavirus/.

2. Lin Y, Hu Z, Zhao Q, Alias H, Danaee M, Wong LP. Understanding COVID-19 vaccine demand and hesitancy: A nationwide online survey in China. PLoS neglected tropical diseases. 2020;14(12):e0008961. Epub 2020/12/18. doi: 10.1371/journal.pntd.0008961. PubMed PMID: 33332359; PubMed Central PMCID: PMCPMC7775119.

3. World Health Organization. Draft landscape and tracker of COVID-19 candidate vaccines: World Health Organization; [cited 2021 26 January]. Available from: https://www.who.int/publications/m/item/draft-landscape-of-covid-19-candidate-vaccines.

4. World Health Organization. WHO issues its first emergency use validation for a COVID-19 vaccine and emphasizes need for equitable global access 2020 [cited 2021 1 February]. Available from: https://www.who.int/news/item/31-12-2020-who-issues-its-first-emergency-use-validation-for-a-covid-19-vaccine-and-emphasizes-need-for-equitable-global-access#:~:text=The%20World%20Health%20Organization%20(WHO,outbreak%20began%20a%20year%20ago.

5. Wang J, Peng Y, Xu H, Cui Z, Williams RO, 3rd. The COVID-19 Vaccine Race: Challenges and Opportunities in Vaccine Formulation. AAPS PharmSciTech. 2020;21(6):225-. doi: 10.1208/s12249-020-01744-7. PubMed PMID: 32761294.

6. Sharma O, Sultan AA, Ding H, Triggle CR. A Review of the Progress and Challenges of Developing a Vaccine for COVID-19. Front Immunol. 2020;11:585354-. doi: 10.3389/fimmu.2020.585354. PubMed PMID: 33163000.

7. Teerawattananon Y, Dabak SV. COVID vaccination logistics: five steps to take now. Nature. 2020;587(7833):194-6. Epub 2020/11/11. doi: 10.1038/d41586-020-03134-2. PubMed PMID: 33168970.

8. Dror AA, Eisenbach N, Taiber S, Morozov NG, Mizrachi M, Zigron A, et al. Vaccine hesitancy: the next challenge in the fight against COVID-19. European Journal of Epidemiology. 2020;35(8):775–9. doi: 10.1007/s10654-020-00671-y.

9. MacDonald NE. Vaccine hesitancy: Definition, scope and determinants. Vaccine. 2015;33(34):4161-4. Epub 2015/04/22. doi: 10.1016/j.vaccine.2015.04.036. PubMed PMID: 25896383.

10. Dubé E, Laberge C, Guay M, Bramadat P, Roy R, Bettinger JA. Vaccine hesitancy: an overview. Human vaccines & immunotherapeutics. 2013;9(8):1763–73.

11. Palamenghi L, Barello S, Boccia S, Graffigna G. Mistrust in biomedical research and vaccine hesitancy: the forefront challenge in the battle against COVID-19 in Italy. European journal of epidemiology. 2020;35(8):785–8.

12. Lazarus JV, Ratzan SC, Palayew A, Gostin LO, Larson HJ, Rabin K, et al. A global survey of potential acceptance of a COVID-19 vaccine. Nature medicine. 2020:1-4. Epub 2020/10/22. doi: 10.1038/s41591-020-1124-9. PubMed PMID: 33082575; PubMed Central PMCID: PMCPMC7573523.

13. Sallam M. COVID-19 vaccine hesitancy worldwide: a systematic review of vaccine acceptance rates. medRxiv. 2021:2020.12. 28.20248950.

14. Gilkey MB, Magnus BE, Reiter PL, McRee A-L, Dempsey AF, Brewer NT. The Vaccination Confidence Scale: a brief measure of parents’ vaccination beliefs. Vaccine. 2014;32(47):6259–65.

15. Opel DJ, Mangione-Smith R, Taylor JA, Korfiatis C, Wiese C, Catz S, et al. Development of a survey to identify vaccine-hesitant parents: the parent attitudes about childhood vaccines survey. Human vaccines. 2011;7(4):419–25.

16. 11. Shapiro GK, Tatar O, Dube E, Amsel R, Knauper B, Naz A, et al. The vaccine hesitancy scale: Psychometric properties and validation. Vaccine. 2018;36(5):660–7.

17. Larson HJ, De Figueiredo A, Xiahong Z, Schulz WS, Verger P, Johnston IG, et al. The state of vaccine confidence 2016: global insights through a 67-country survey. EBioMedicine. 2016;12:295–301.

18. Betsch C, Schmid P, Heinemeier D, Korn L, Holtmann C, Böhm R. Beyond confidence: Development of a measure assessing the 5C psychological antecedents of vaccination. PloS one. 2018;13(12):e0208601–e. doi: 10.1371/journal.pone.0208601. PubMed PMID: 30532274.

19. Betsch C, Schmid P, Heinemeier D, Korn L, Holtmann C, Böhm R. Beyond confidence: Development of a measure assessing the 5C psychological antecedents of vaccination. PloS one. 2018;13(12):e0208601.

20. Sallam M, Dababseh D, Eid H, Al-Mahzoum K, Al-Haidar A, Taim D, et al. High Rates of COVID-19 Vaccine Hesitancy and Its Association with Conspiracy Beliefs: A Study in Jordan and Kuwait among Other Arab Countries. Vaccines. 2021;9(1):42.

21. Al-Mohaithef M, Padhi BK. Determinants of COVID-19 vaccine acceptance in Saudi Arabia: a web-based national survey. Journal of multidisciplinary healthcare. 2020;13:1657.

22. Brewer NT, Cuite CL, Herrington JE, Weinstein ND. Risk compensation and vaccination: can getting vaccinated cause people to engage in risky behaviors? Annals of behavioral medicine : a publication of the Society of Behavioral Medicine. 2007;34(1):95-9. Epub 2007/08/11. doi: 10.1007/bf02879925. PubMed PMID: 17688401.

23. Fine P, Eames K, Heymann DL. “Herd Immunity”: A Rough Guide. Clinical Infectious Diseases. 2011;52(7):911–6. doi: 10.1093/cid/cir007.

24. Betsch C, Bach Habersaat K, Deshevoi S, Heinemeier D, Briko N, Kostenko N, et al. Sample study protocol for adapting and translating the 5C scale to assess the psychological antecedents of vaccination. BMJ Open. 2020;10(3):e034869. doi: 10.1136/bmjopen-2019-034869.

25. Pedhazur EJ, Kerlinger FN. Multiple regression in behavioral research: Holt, Rinehart, and Winston; 1982.

26. Soper DS. A-priori sample size calculator for structural equation models [Software]. Recuperado de http://www.danielsopercom/statcalc. 2017.

27. Cronbach LJ. Coefficient alpha and the internal structure of tests. psychometrika. 1951;16(3):297–334.

28. Boateng GO, Neilands TB, Frongillo EA, Melgar-Quiñonez HR, Young SL. Best Practices for Developing and Validating Scales for Health, Social, and Behavioral Research: A Primer. Frontiers in public health. 2018;6:149. Epub 2018/06/27. doi: 10.3389/fpubh.2018.00149. PubMed PMID: 29942800; PubMed Central PMCID: PMCPMC6004510.

29. Drost EA. Validity and reliability in social science research. Education Research and perspectives. 2011;38(1):105.

30. Ockey GJ. Exploratory factor analysis and structural equation modeling. The companion to language assessment. 2013;3:1224–44.

31. Samuels P. Advice on exploratory factor analysis. 2017.

32. Field AP. Discovering statistics using SPSS for Windows: Advanced techniques for the beginner: Sage; 2009.

33. Field A. Discovering statistics using IBM SPSS statistics: sage; 2013.

34. Fabrigar LR, Wegener DT, MacCallum RC, Strahan EJ. Evaluating the use of exploratory factor analysis in psychological research. Psychological methods. 1999;4(3):272.

35. Brown TA, Moore MT. Confirmatory factor analysis. Handbook of structural equation modeling. 2012:361–79.

36. Marsh HW, Balla JR, McDonald RP. Goodness-of-fit indexes in confirmatory factor analysis: The effect of sample size. Psychological bulletin. 1988;103(3):391.

37. Fornell C, Larcker DF. Evaluating structural equation models with unobservable variables and measurement error. Journal of marketing research. 1981;18(1):39–50.

38. Rose S. International ethical guidelines for epidemiological studies: by the Council for International Organizations of Medical Sciences (CIOMS). Oxford University Press; 2009.

39. Hilton A, Skrutkowski M. Translating instruments into other languages: development and testing processes. Cancer nursing. 2002;25(1):1–7.

40. Cheung GW, Wang C, editors. Current approaches for assessing convergent and discriminant validity with SEM: Issues and solutions. Academy of management proceedings; 2017: Academy of Management Briarcliff Manor, NY 10510.

