## Supplementary table 1 for "Arabic validation and cross-cultural adaptation of the 5C scale for assessment of COVID-19 vaccines psychological antecedents"

**Table 1: Inter-item correlations of the Arabic version of the 5C scale**

| **Confidence** | | | **Complacency** | | | **Constraints** | | | **Calculation** | | | **Collective responsibility** | | |
| --- | --- | --- | --- | --- | --- | --- | --- | --- | --- | --- | --- | --- | --- | --- |
|  | Q2 | Q3 |  | Q5 | Q6 |  | Q8 | Q9 |  | Q11 | Q12 |  | Q14 | Q15 |
| Q1 | 0.81(*P*<0.001) | 0.58(*P*<0.001) | Q4 | 0.50(*P*<0.001) | 0.47(*P*<0.001) | Q7 | 0.35(*P*<0.001) | 0.28(*P*<0.001) | Q10 | 0.58(*P*<0.001) | 0.46(*P*<0.001) | Q13 | 0.56(*P*<0.001) | 0.58(*P*<0.001) |
| Q2 |  | 0.51(*P*<0.001) | Q5 |  | 0.40(*P*<0.001) | Q8 |  | 0.53(*P*<0.001) | Q11 |  | 0.59(*P*<0.001) | Q14 |  | 0.74(*P*<0.001) |
